# Antimicrobial Resistance Rates in Gram-positive Uropathogens in Duhok city, Kurdistan Region of Iraq

**DOI:** 10.1101/2023.02.26.23286459

**Authors:** Alan Ali Mohamed

**Affiliations:** Department of Medical Health Sciences, College of Medicine, University of Zakho, Kurdistan Region, Iraq

**Keywords:** Urinary Tract Infection, Antimicrobial resistance, Uropathogens

## Abstract

**Background:** Urinary tract infections (UTIs) are among the most common infections world-wide. Antibiotic resistance is an important medical problem because there is an increasing trend of antibiotic resistance worldwide making it harder to eliminate uropathogens. Antibiotic resistance shows a geographical variation. Hence, local studies are necessary to determine prevalence of uroptahogens among UTI patients. Thus, this study was conducted to determine prevalence of uropathogens among UTI patients and their antimicrobial susceptibility pattern from the data of 12 years period from 2010 to 2022 in Duhok Province, Kurdistan Region, Iraq

**Materials and methods:** This study was conducted by retrieving 12-year laboratory records between 2010–2022. data were collected from Azadi teaching hospital in Duhok city in Kurdistan region in northern Iraq. uropathogen species were identified by routine laboratory methods. Antimicrobial sensitivity testing was performed manually and by Vitek-2 automated susceptibility system.

**Results:** The results of gram-positive urine pathogens of 249 patients showed Staphylococcus haemolyticus (20.9%) was the most common isolated pathogen, followed by staphylococcus aureus (7.6%), Streptococcus Agalactiae (6.4%) and Enterococcus spp. (6%). S. haemolyticus had highest resistance to nitrofurantoin (61.5%) and lowest amikacin (34.3%). S. aureus showed highest resistance to penicillin 73.3% and highest sensitivity to amikacin (78.9%).in our study, Streptococcus agalactiae had the highest sensitivity to vancomycin (%87.5). 93.3% of enterococcus spp. were resistant to tetracycline.

**Conclusion:** UTIs are more common in female than in male and coagulase-negative Staph species (Staphylococcus spp) are the most commonly isolated pathogens. This study found antimicrobial resistance to commonly prescribed antibiotics are high. Hence, an urgent plan to control antimicrobial resistance is necessary in our area.

## Introduction

Urinary tract infections (UTIs) are among the most common bacterial infections with an estimation of more than 150 million infections, annually world-wide [1]. UTIs cause various clinical sign and symptoms including dysuria, nocturia, polyuria, pelvic pain, pyrexia, chills, nausea and vomiting [2, 3]. Escherichia coli is the most common bacterial uropathogen causing UTIs (which is responsible of 75% to 90% cases of UTIs) followed by Klebsiella pneumoniae, Citrobacter species, Staphylococcus aureus, Staphylococcus saprophyticus and Pseudomonas aeruginosa [2, 4, 5]. The infection is more common in females due to certain anatomical differences with an increased risk of infection among individuals with frequent sexual intercourse, infants, elderly, abnormalities of urinary tract, low socio-economic status, pregnant women, diabetic and spinal cord injury patients [6, 7]. Approximately, %50 of women will develop UTIs once in their life time and between 20% to 40% of them experience recurrent episodes of UTIs, Whilst 20% of UTI cases are men [8]. Treatment of UTIs varies according to age, sex, underlying disease, infective organism and involvement of upper or lower urinary tract [9, 10].

Antibiotic resistance rate changes according to geographical areas due to presence of different strains in different regions of the world and variation in antibiotic use [11]. Resistance pattern of pathogens to antibiotics are continuously changing [9, 12-14]. Antibiotic resistance is an important medical problem and one of the World Health Organization (WHO) priorities because there is an increasing trend of antibiotic resistance world-wide making it harder to eliminate uropathogens from urinary tract which end up with longer illness and mortality rates [15, 16]. Antimicrobial resistance also causes a financial burden due to extra laboratory tests and usage of more expensive drugs is needed for treatment of resistant strains [4]. As in majority of infectious diseases like UTIs, Empirical treatment is given before causative organisms and their susceptibility pattern are obtained, therefore physicians must have a good understanding of common etiological organisms of UTIs and their susceptibility pattern to determine the appropriate treatment based on the most likely pathogen and sensitivity of it in a geographical area [17-19]. periodic monitoring of change in sensitivity of pathogens to antibiotics is vital [20]. Antibiotic sensitivity patterns have been studied in Kurdistan region of Iraq [21-26]. These studies showed that the antibiotic resistance rates are high [27-29]. Hence, this study is conducted to determine prevalence of Gram-positive uropathogens among UTI patients, and to assess their antimicrobial susceptibility pattern to commonly used antimicrobial agent to help with doctors to determine appropriate empirical treatment as well as to detect any possible trend of change in sensitivity during a 12 years period from 2010 to 2022 in Duhok Province, Kurdistan Region, Iraq

## Materials and methods

### Study setting

This was a retrospective cross-sectional study which was conducted by retrieving 12-year laboratory records (2010–2022) in Duhok city, Kurdistan Region of Iraq. The study was conducted between 1st of September, 2022 - 1st of March 2023. data were collected from Azadi teaching hospital (tertiary hospital) in Duhok city in Kurdistan region in north Iraq which serves more than 2 million people in Duhok province and around neighboring regions. A number of UTI suspected patient urine culture results had been recruited in the study from hospital’s microbiology laboratory logbook of twelve years (2010 – 2022) which is a paper containing patient’s name, gender, identified organism and antibiotic sensitivity of identified organism.

### Antibiotic Sensitivity testing and sample collection

urine samples of UTI suspected patients were collected by clean catch midstream method. Collected samples were inoculated on blood and MacConkey agar and incubated for 24 hours at 37° C. bacterial colonies were initially classified by gram staining and determined according to standard cultures and their biochemical characteristics. Antimicrobial sensitivity testing was performed manually and by Vitek-2 automated susceptibility system according to manufacturer’s instructions.

### Ethics

The study protocol was approved by the ethics committee at the University of Duhok, college of medicine, Kurdistan Region, Iraq.

## Results

### Patient’s characteristics

A total of 249 patients within different age groups had been recruited in this study. 77.5% (193) of the patients were female, while 22.5% (56) of the patients were male. Female to male ratio was 3.4:1 (**Table 1**).

**Table 1:** distribution of urine pathogens by gender.

| Organism | female No. (%) | male No. (%) | total No. (%) |
| --- | --- | --- | --- |
| **staphylococcus Haemolyticus** | 35 (18.1) | 17 (30.4) | 52 (20.9) |
| **staphylococcus aureus** | 14 (7.3) | 5 (8.9) | 19 (7.6) |
| **Streptococcus Agalactiae** | 15 (7.8) | 1 (1.8) | 16 (6.4) |
| **Enterococcus spp.** | 13 (6.7) | 2 (3.6) | 15 (6) |
| **All other staphylococcus spp.** | 85 (44.1) | 21 (37.5) | 106 (42.6) |
| **all other streptococcus spp.** | 31 (16) | 10 (17.9) | 41(16.5) |
| **Total** | 193 (100) | 56 (100) | 249 (100) |

\*Data had been reported as numbers of isolated pathogens and percentages (within brackets) of total isolated gram-positive uropathogens

### Microorganism characteristic

The results of gram-positive urine pathogens of our study showed that Staphylococcus haemolyticus had the highest infection rate (20.9%), followed by staphylococcus aureus (7.6%). Streptococcus Agalactiae were the third most common pathogen (6.4%). A significant proportion of infections were by coagulase-negative Staph species (Staphylococcus spp) (42.6%) (excluding Staph. Haemolyticus) and other Streptococcus spp. (16.5%) (excluding streptococcus agalactiae) (**Table1**).

Regarding antimicrobial sensitivity patterns, staphylococcus haemolyticus (which is the commonest isolated urine pathogen in this study) showed higher resistance rate to nitrofurantoin (61.5%), Tetracycline (59.6%), penicillin (53.8%) and lower resistance rate to gentamicin (46.2%), ciprofloxacin (44.2%), trimethoprim/sulfamethoxazole(42.3%), amikacin (34.6%) and vancomycin (34.6%) (**table 2**). In the terms of sensitivity pattern of Staphylococcus aureus isolates, the data showed 73.3% and 63.2% of isolates were resistant to penicillin and nitrofurantoin respectively, while 78.9% of isolates were sensitive to amikacin (**table 2**). Additionally, %87.5 of Streptococcus agalactiae isolates were found to be sensitive to vancomycin (**table 2**). 93.3% of enterococcus spp. were resistant to tetracycline in our data (**table 2**). In this study, All Staph. spp. showed highest sensitivity to amikacin, vancomycin and nitrofurantoin (58.5%), while showed highest resistance rate to penicillin (64.2%) **(table 2**). Additionally, %73.2 of all other strept. Spp. were found to be resistant to gentamicin (**table 2**).

**Table 2:** Antimicrobial Susceptibility Patterns of Isolated pathogens.

| Organism (number of isolates) | Sensitivity pattern (%) | Antimicrobial Agent |  |  |  |  |  |  |  |
| --- | --- | --- | --- | --- | --- | --- | --- | --- | --- |
|  |  | Penicillin or benzylpenicillin | Gentamicin | Amikacin | Ciprofloxacin | Vancomycin | Tetracycline | Nitrofurantoin | trimethoprim/sulfa methoxazole |
| **Staph. aureus (19)** | **Sensitive** | 15.8 | 42.1 | 78.9 | 36.8 | 47.4 | 15.8 | 31.6 | 31.6 |
|  | **Intermediate** | 10.5 | 21.1 | 5.3 | 10.5 | 15.8 | 26.3 | 5.3 | 21.1 |
|  | **Resistant** | 73.7 | 36.8 | 15.8 | 52.6 | 36.8 | 57.9 | 63.2 | 47.4 |
| **Staph. Hemolyticus (52)** | **Sensitive** | 34.6 | 40.4 | 50 | 46.2 | 63.5 | 21.2 | 32.7 | 51.9 |
|  | **Intermediate** | 11.5 | 13.5 | 15.4 | 9.6 | 1.9 | 19.2 | 5.8 | 5.8 |
|  | **Resistant** | 53.8 | 46.2 | 34.6 | 44.2 | 34.6 | 59.6 | 61.5 | 42.3 |
| **all other staph. spp. (106)** | **Sensitive** | 22.6 | 36.8 | 58.5 | 56.6 | 58.5 | 37.7 | 58.5 | 53.8 |
|  | **Intermediate** | 13.2 | 16 | 19.8 | 11.3 | 24.5 | 26.4 | 8.5 | 17.9 |
|  | **Resistant** | 64.2 | 47.2 | 21.7 | 32.1 | 17 | 35.8 | 33 | 28.3 |
| **Enterococcus spp. (15)** | **Sensitive** | 60 | 60 | 53.3 | 53.3 | 53.3 | 6.7 | 33.3 | 40 |
|  | **Intermediate** | 0 | 6.7 | 6.7 | 13.3 | 13.3 | 0 | 6.7 | 13.3 |
|  | **Resistant** | 40 | 33.3 | 40 | 33.3 | 33.3 | 93.3 | 60 | 46.7 |
| **Strept. Agalactiae (16)** | **Sensitive** | 56.3 | 56.3 | 62.5 | 56.3 | 87.5 | 37.5 | 50 | 56.3 |
|  | **Intermediate** | 0 | 0 | 6.3 | 6.3 | 12.5 | 6.3 | 6.3 | 0 |
|  | **Resistant** | 43.8 | 43.8 | 31.3 | 37.5 | 0 | 56.3 | 43.8 | 43.8 |
| **all other strept. Spp. (41)** | **Sensitive** | 41.5 | 24.4 | 31.7 | 51.2 | 58.5 | 48.8 | 43.9 | 48.8 |
|  | **Intermediate** | 7.3 | 2.4 | 17.1 | 7.3 | 26.8 | 17.1 | 19.5 | 7.3 |
|  | **Resistant** | 51.2 | 73.2 | 51.2 | 41.5 | 14.6 | 34.1 | 36.6 | 43.9 |

#### Infection In female

Regarding distribution of isolated urine pathogens among female individuals, Staphylococcus haemolyticus had the highest infection rate (18.1%), followed by Streptococcus Agalactiae (7.8%) with staphylococcus aureus being the third most common isolated pathogen (7.3%) **(table 3)** .A significant proportion of infections were by other Staphylococcus spp. (44.1%) and other Streptococcus spp. (16%) (**table 3**).

**Table 3:** Antimicrobial Susceptibility Patterns of Isolated pathogens of female participants.

| Organism (number of isolates) | Sensitivity pattern (%) | Antimicrobial Agent |  |  |  |  |  |  |  |
| --- | --- | --- | --- | --- | --- | --- | --- | --- | --- |
|  |  | Penicillin or benzylpenicillin | Gentamicin | Amikacin | Ciprofloxacin | Vancomycin | Tetracycline | Nitrofurantoin | trimethoprim/sulfamethoxazole |
| Staph. Aureus (14) | Sensitive | 14.3 | 42.9 | 78.6 | 50 | 42.9 | 14.3 | 21.4 | 35.7 |
|  | Intermediate | 7.1 | 28.6 | 7.1 | 7.1 | 21.4 | 21.4 | 7.1 | 21.4 |
|  | Resistant | 78.6 | 28.6 | 14.3 | 42.9 | 35.7 | 64.3 | 71.4 | 42.9 |
| Staph. haemolyticus (35) | Sensitive | 31.4 | 40 | 48.6 | 51.4 | 57.1 | 22.9 | 31.4 | 48.6 |
|  | Intermediate | 14.3 | 17.1 | 17.1 | 11.4 | 0 | 20 | 5.7 | 5.7 |
|  | Resistant | 54.3 | 42.9 | 34.3 | 37.1 | 42.9 | 57.1 | 62.9 | 45.7 |
| all other staph. spp. (85) | Sensitive | 18.4 | 39.1 | 64.4 | 56.3 | 56.3 | 40.2 | 60.9 | 54 |
|  | Intermediate | 13.8 | 18.4 | 16.1 | 11.5 | 25.3 | 23 | 8 | 17.2 |
|  | Resistant | 67.8 | 42.5 | 19.5 | 32.2 | 18.4 | 36.8 | 31 | 28.7 |
| Enterococcus spp. (13) | Sensitive | 53.8 | 53.8 | 46.2 | 53.8 | 61.5 | 7.7 | 38.5 | 46.2 |
|  | Intermediate | 0 | 7.7 | 7.7 | 15.4 | 7.7 | 0 | 7.7 | 15.4 |
|  | Resistant | 46.2 | 38.5 | 46.2 | 30.8 | 30.8 | 92.3 | 53.8 | 38.5 |
| Strept. Agalactiae (15) | Sensitive | 60 | 60 | 66.7 | 60 | 86.7 | 40 | 46.7 | 53.3 |
|  | Intermediate | 0 | 0 | 6.7 | 0 | 13.3 | 0 | 6.7 | 0 |
|  | Resistant | 40 | 40 | 26.7 | 40 | 0 | 60 | 46.7 | 46.7 |
| all other strept. Spp. (31) | Sensitive | 41.5 | 24.4 | 31.7 | 51.2 | 58.5 | 48.8 | 43.9 | 48.8 |
|  | Intermediate | 7.3 | 2.4 | 17.1 | 7.3 | 26.8 | 17.1 | 19.5 | 7.3 |
|  | Resistant | 51.2 | 73.2 | 51.2 | 41.5 | 14.6 | 34.1 | 36.6 | 43.9 |

Regarding sensitivity patterns, staphylococcus haemolyticus was found to be have higher resistance rate to nitrofurantoin (62.9%), Tetracycline (57.1%), penicillin (54.3%) and lower resistance rate to amikacin (34.3%) ciprofloxacin (37.1%), gentamicin (42.9%), vancomycin (42.9%) and trimethoprim/sulfamethoxazole(45.7%) (**table 3**). Streptococcus Agalactiae were found to be the second most common organism among females and had the highest sensitivity to vancomycin (86.7%), while showing the highest resistance to tetracycline (60%).Staph. aureus showed the highest resistance to Penicillin (78.6%) and nitrofurantoin (71.4%), whilst having highest sensitivity for amikacin (78.6%) (**table 3**). In the terms of sensitivity patterns of enterococcus spp., it was revealed that 92.3% of isolates were resistance to tetracycline, while 61.5% were sensitive to vancomycin (**table 3**).

#### Infection in male

Regarding distribution of isolated urine pathogens among male participants, Staphylococcus haemolyticus had the highest infection rate (30.4%), followed by staphylococcus aureus (8.9%), while Streptococcus Agalactiae showed the lowest rate (1.8%) **(table 4)**. A significant proportion of infections were by other Staphylococcus spp. (37.5%) and other Streptococcus spp. (17.9%) (**table 4**). Regarding sensitivity patterns of males, staphylococcus haemolyticus was found to have higher resistance rate to Tetracycline (57.1%), nitrofurantoin (62.9%), penicillin (52.9%), gentamicin (52.9%), While showing lower resistance rate to amikacin(35.3%), trimethoprim/sulfamethoxazole (35.3%) vancomycin (17.6%) (**table 4**). In our data, Staph. aureus isolates were the most resistant to ciprofloxacin(80%), while being the most sensitive to amikacin (80%). Additionally, All other strept. Spp. showed 77.8% resistance rate to gentamicin. **(table 4)**

**Table 4:**
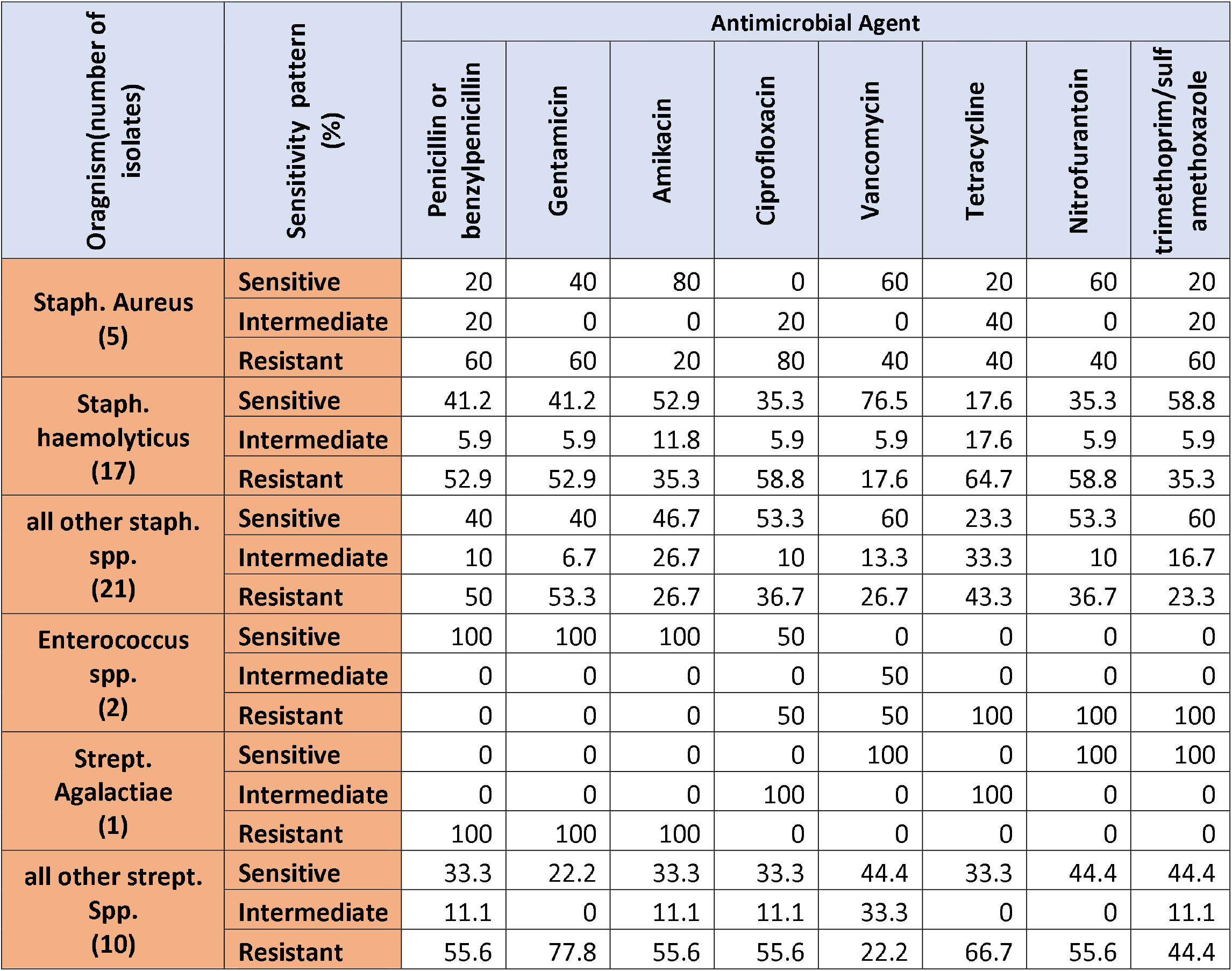
Antimicrobial Susceptibility Patterns of Isolated pathogens of male participants.

| Organism(number of isolates) | Sensitivity pattern (%) | Antimicrobial Agent |  |  |  |  |  |  |  |
| --- | --- | --- | --- | --- | --- | --- | --- | --- | --- |
|  |  | Penicillin or benzylpenicillin | Gentamicin | Amikacin | Ciprofloxacin | Vancomycin | Tetracycline | Nitrofurantoin | trimethoprim/sulfamethoxazole |
| Staph. Aureus (5) | Sensitive | 20 | 40 | 80 | 0 | 60 | 20 | 60 | 20 |
|  | Intermediate | 20 | 0 | 0 | 20 | 0 | 40 | 0 | 20 |
|  | Resistant | 60 | 60 | 20 | 80 | 40 | 40 | 40 | 60 |
| Staph. haemolyticus (17) | Sensitive | 41.2 | 41.2 | 52.9 | 35.3 | 76.5 | 17.6 | 35.3 | 58.8 |
|  | Intermediate | 5.9 | 5.9 | 11.8 | 5.9 | 5.9 | 17.6 | 5.9 | 5.9 |
|  | Resistant | 52.9 | 52.9 | 35.3 | 58.8 | 17.6 | 64.7 | 58.8 | 35.3 |
| all other staph. spp. (21) | Sensitive | 40 | 40 | 46.7 | 53.3 | 60 | 23.3 | 53.3 | 60 |
|  | Intermediate | 10 | 6.7 | 26.7 | 10 | 13.3 | 33.3 | 10 | 16.7 |
|  | Resistant | 50 | 53.3 | 26.7 | 36.7 | 26.7 | 43.3 | 36.7 | 23.3 |
| Enterococcus spp. (2) | Sensitive | 100 | 100 | 100 | 50 | 0 | 0 | 0 | 0 |
|  | Intermediate | 0 | 0 | 0 | 0 | 50 | 0 | 0 | 0 |
|  | Resistant | 0 | 0 | 0 | 50 | 50 | 100 | 100 | 100 |
| Strept. Agalactiae (1) | Sensitive | 0 | 0 | 0 | 0 | 100 | 0 | 100 | 100 |
|  | Intermediate | 0 | 0 | 0 | 100 | 0 | 100 | 0 | 0 |
|  | Resistant | 100 | 100 | 100 | 0 | 0 | 0 | 0 | 0 |
| all other strept. Spp. (10) | Sensitive | 33.3 | 22.2 | 33.3 | 33.3 | 44.4 | 33.3 | 44.4 | 44.4 |
|  | Intermediate | 11.1 | 0 | 11.1 | 11.1 | 33.3 | 0 | 0 | 11.1 |
|  | Resistant | 55.6 | 77.8 | 55.6 | 55.6 | 22.2 | 66.7 | 55.6 | 44.4 |

## Discussion

UTIs are among the most common infection world-wide. due to development of urosepsis, renal scarring or progressive kidney disease, UTIs pose a serious health concern. UTIs are treated by antibiotics and empirical treatment by appropriate antimicrobial agents is an important factor in prognosis of the disease[20]. antimicrobial resistance to shows a geographical variation [30, 31]. Hence periodic monitoring of local antibiotic sensitivity patterns to urine pathogens by studies like this is necessary [32]. In this study, it was found that UTIs are significantly more common in female than in male with 77.5% and 22.5% of our cases being female and male respectively. This was in agreement with previous studies which revealed UTIs are more common in female than in male [8]. The reason behind higher infection rates among females is due to specific anatomical and physiological differences with estimations showing half of females will develop UTI at least once in their lifetime [3, 8, 33]. In addition, sexual intercourse is regarded as a risk factor for developing UTI among females [34].

Among the total of 249 cases of gram-positive pathogens in this study, staphylococcus spp. were the most commonly isolated bacteria which accounted for 71.1% of isolates, while Staphylococcus haemolyticus (20.9%) most commonly isolated specie with Staphylococcus aureus(7.6%) being the second most commonly isolated pathogen in our area (**table 1**).This data is in agreement with other studies in Iraq [32, 35, 36]. In another study conducted Iran showed that Staphylococcus spp. as the most common etiological gram-positive urine pathogens [31, 34, 37].

Our findings demonstrated that staphylococcus haemolyticus isolates were the most sensitive to vancomycin and amikacin while being the most resistance to nitrofurantoin and tetracycline. These data were similar to previous reports conducted in Iraq [32, 35, 38]. A significant difference of overall resistance rate of S.haemolyticus to antimicrobial agents in our study between male and female patients has not been found. In present study, S. aureus showed high resistance penicillin (73.3%) and nitrofurantoin (63.2%) and high sensitivity to amikacin (78.9%). other reports in Iraq revealed 100% penicillin resistance which is a higher resistance rate than our findings and 20% resistance to nitrofurantoin which was lower than our findings [35].

Another study in our locality showed high sensitivity of S. aureus to most of the commonly used antibiotics[36]. In addition, These our results were similar to what was found in a study in Nepal in 2018 which found high sensitivity of this organism to amikacin and high resistance to penicillin with the exception of nitrofurantoin being highly sensitive in the study[39]. The reason behind nitrofurantoin sensitivity difference may be due to sample size or may be due to this drug is more commonly being used in our community, therefore caused higher resistance rate. In the present study a significant difference of resistance rates to S. aureus between males and females had not been recorded.

In this study, Streptococcus agalactiae (also known as Group B Streptococcus(GBS)) showed the highest sensitivity to vancomycin with 87.5%, while had the highest resistance to tetracycline with 56.3%. these findings were comparable to what was found in our locality [32, 35]. In addition, A significant difference has been recorded in frequency of Streptococcus agalactiae in relation to gender in which female had significantly higher infection rates than males with the rates of 7.8% to 1.8% respectively (**table 1**). The difference was also observed in a study in turkey [40]. According to the Centers for Disease Control and Prevention (CDC), approximately 25% of pregnant women are colonized with GBS. Unfortunately, history of pregnancy had not been obtained in the current study.

In the present study, Enterococcus spp. were accounted for a higher percentage in females than in males. Regarding sensitivity patterns of Enterococcus spp. in our study, these organisms were the resistant to tetracycline (93.3%), with lowest resistance to gentamicin, vancomycin and ciprofloxacin with rates of 33.3%. previous international studies had showed different results than from ours [39, 41]. This may be explained by low sample size of enterococcus spp. in the current study.

Regrettably, it has been determined that physicians possess inadequate knowledge regarding antibiotic sensitivity patterns [42]. Furthermore, during the COVID-19 pandemic, antibiotics were utilized indiscriminately [43, 44], which may have contributed to an increase in resistance rates. Consequently, an immediate strategy is imperative to counteract the emergence of antibiotic resistance.

## Conclusion

From this study, we’ve concluded that UTIs are more common in female than in male, and Staphylococcus species are the most common etiological gram-positive organisms causing UTI. This study revealed high resistance rate among commonly used antimicrobial agents. Urgent plan has to be made to stop increasing trends of antimicrobial resistance. Furthermore, more studies should be conducted to further evaluate local resistance of commonly prescribed antibiotics for a better establishment of empirical treatment of UTIs.

## Data Availability

All data produced in the present work are contained in the manuscript

